# Preventive chemotherapy coverage against Soil-transmitted helminthiases among school age children in vertical versus integrated treatment approaches: Implications from coverage validation survey in Ethiopia

**DOI:** 10.1101/2020.03.18.20038620

**Authors:** Mekuria Asfaw, Zerihun Zerdo, Chuchu Churko, Fikre Seife, Manaye Yihune, Yilma Chisha, Abinet Teshome, Birhanu Getachew, Nebiyu Negussu

## Abstract

**Background:** Soil-transmitted helminths (STH) are widely distributed in Ethiopia with highest prevalence and burden. Since 2015 the country launched national deworming programme to control STH associated morbidity using mass treatment with Albendazole/Mebendazole. Data on routine coverage of Preventive chemotherapy (PC) are available at different level of the health system, however in some circumstances these reports are unreliable and evidence is lacking on validated treatment coverage against STH.

**Methodology:** A community-based cross-sectional study was conducted in ten districts of Ethiopia; from January to April 2019. A total of 8154 SAC (4100 males and 4054 females) were participated from randomly selected households. Data were analysed using SPSS software (IBM, version 25); then di-aggregated by gender, age and school attendance and presented in tables and graphs.

**Principal findings:** Albendazole/Mebendazole mass treatment coverage against STH among school-age children was found to be 71%. In vertical (school-based) treatment approach, 4822(68.4%) were treated; whereas in integrated (community directed) approach, 963(86.9 %) were treated. The treatment coverage among males was 2948(71.9%), while among females it was 2837(70%). Based on age the treatment coverage in the age group 10-14 years was 77%; which is higher than the coverage in age group 5-9 years was 64.4%. In addition, the treatment coverage in school attendant was 81%; which is higher than coverage non-enrolled children (28%). The main deworming site was school, 5223(91%). Moreover, the main reported reasons for not swallowing drugs were not attending school, 422(19.75%) and drugs were not given, 397(18.6%).

**Concussions/significance:** Albendazole/Mebendazole mass treatment coverage against soil-transmitted helminths among school-age children was below the WHO recommendation (75%). Operational research is required to identify barriers for low coverage of ALB/MBD among children who are non-enrolled school-age children. Further, a call for action is required from different stakeholders to improve treatment coverage of ALB/MBD.

**Author Summary:** Neglected Tropical Diseases (NTDs) are a group of communicable diseases, which inequitably affect the world’s poorest, marginalized, voiceless and powerless people. It is prevalent in areas with unsafe water, poor sanitation and hygiene. STH (ascariasis, hookworm, and trichuriasis), intestinal worms, are one of the common NTDs which are transmitted through contact with soil contaminated with human feces.

Globally, more than 4 billion people are at risk for STH and with over 1 billion are already infected. In Ethiopia, about 81 million people are at risk for STH infection. School age children and pre-school age children are highly affected by the diseases, and it causes anemia, vitamin A deficiency, stunting, malnutrition, impaired development, and intestinal obstruction.

As one of NTDS, ending STH can contribute to Ethiopia be on track of attaining universal health coverage (UHC). STH can be controlled, possibly eliminated by combined interventions of preventive chemotherapy with improved water, sanitation and hygiene (WASH). In Ethiopia, though remarkable attainments are made so far on controlling STH morbidity through mapping and scaling-up mass drug administration, still more work is required to increase coverage of PC and integration of PC with WASH to meet the national objectives of elimination or control of STH.

## Introduction

Neglected tropical diseases (NTDs) are a group of diseases mainly affect poorest and marginalized people living in rural and urban areas, primarily in tropical and subtropical areas [1, 2]. Soil-transmitted helminthiasis (STH), one of the major Neglected Tropical Diseases (NTDs), are a group of intestinal parasites consisting of Ascaris lumbricoides (roundworms), Trichuris trichiura (whipworms), and Hook worm, are transmitted by faecal contamination of soil, and prevalent in areas with lack of improved water, sanitation and hygiene [3-4].

Globally, about two billion people are being estimated in developing countries that are infected with one or more species of helminths [5]. In 2017, 1.9 million disability-adjusted life years (DALYs) associated with STH infection worldwide are estimated [6, 7]. Ethiopia has wide distribution, highest prevalence and burden of STH, which causes 873,500 disability-adjusted life-years (DALYs), annually which represent 1.9% of the total DALYs lost due to all causes. In the country, about 81 million people are living in STH endemic areas, of which 25.3 million are school-aged children [8, 9]. STH infections with moderate and heavy intensity are associated with anemia, malnutrition, educational loss, and cognitive deficits [10-12].

The World Health Organization (WHO) recommends preventive chemotherapy (PC) with Albendazole (ALB) or Mebendazole (MBD) to control STH-related morbidity in combination with other recommended interventions such as health education and improvement of interventions such as hygiene, water and sanitation [5, 13]. WHO sets a target of regular provision of anthelminthic treatment to cover at least 75% of school age children in endemic area [14]. In 2017, more than 500 million SAC (69% of total SAC in need) received PC for STH globally, with 73% of implementation units reaching 75% effective coverage [15]. More than 596 million SAC in 101 countries were estimated in need of PC for STH in 2017; 61 countries submitted reports on treatment [4].

Ending NTDs by 2030 is targeted in the Sustainable Development Goal 3 (target 3.3); with a focus of equity and Universal Health Coverage (UHC) [16]. Thus, addressing NTDs contributes for achievement of the vision of universal health coverage, which means that all individuals and communities who are in need of the health services will be addressed without suffering financial hardship [17].

Starting from the time when the first World health organization (WHO) road map for the prevention and control of NTDs was issued in 2012, significant progress has been made in terms of controlling STH [2]. In line with the WHO goal, Ethiopia sets a target to achieve a minimum 75% coverage of Albendazole or Mebendazole in school and pre-school age children that should be achieved by the country program by 2020 [9].

In fact, routine coverage data are available at different level of Ethiopian health system. However, evidence is lacking on validated treatment coverage Albendazole or Mebendazole against STH among school age children. These data are required for the following programmatic purposes: First, determine treatment coverage of Albendazole or Mebendazole in school and pre-school age children in vertical and integrated treatment approaches to improve program planning of PC. Second, generate treatment coverage data, dis-aggregated by gender, sex and school attendance. Third, identify the reasons for why some individuals do not receive drugs. Additionally, it can also be used to identify main deworming sites of drug distribution.

## Materials and Methods

### Study setting

Ethiopia is the second largest country by population in sub-Saharan Africa, and is home to over 85 million people in need of preventive treatment, has a much higher burden of these diseases than in other countries of sub-Saharan Africa. Since the launch of the first national master plan for NTDs in 2013, the Government of Ethiopia has been liaising with WHO and other partners to achieve its objectives [9, 18]. **Fig 1** shows the area where study was conducted.

**Fig 1.**
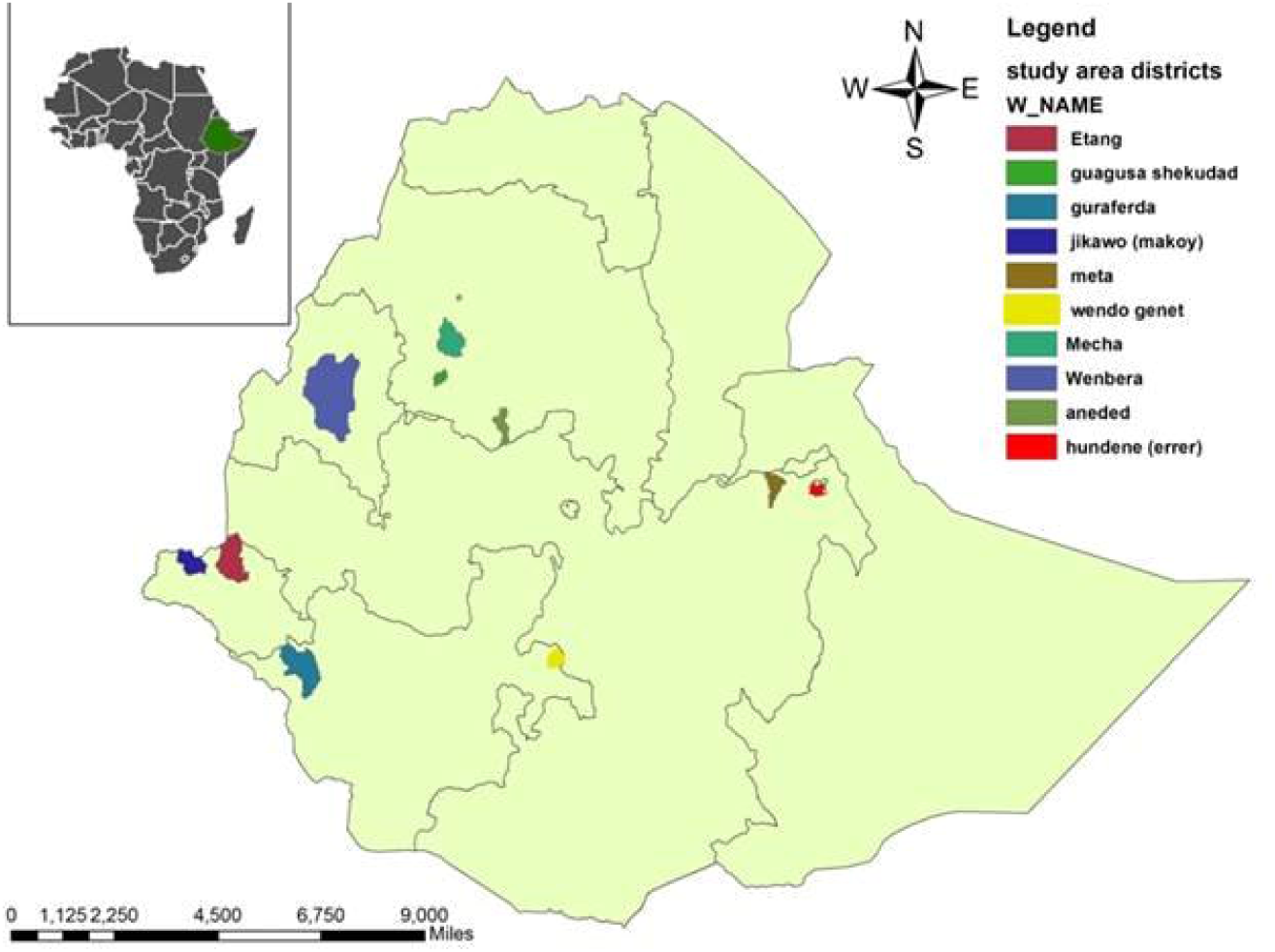
Map of study area for coverage of Albendazole/Mebendazole in Ethiopia, 2019.

### Study design and population

We conducted a community-based cross-sectional study in ten districts of Ethiopia (selected from sentinel site of Federal Ministry of Health); from January to April 2019. All selected school age children who were available during the survey were included in the study.

### Sample size and Sampling technique

A total of 30 segments were randomly selected from each district. The name of kebeles, number of households and the number of segments to be involved were predetermined using survey builder method before going to field for data collection. From each selected segment, at least 16 randomly selected households were included in the survey. From each selected household, all SAC were involved in the survey. Segment means group of 50 households. The sampling is performed using the <u>WHO Coverage Evaluation Guidelines for Preventive Chemotherapy</u> [16].

### Data collection and Statistical analysis

The data were collected using Survey CTO software using smartphones. Data were cleaned and analyzed using an Excel spread sheet and SPSS software (IBM, version 25). Then, data were dis-aggregated by gender, age and school attendance; presented in tables and figures.

### Measurements

Coverage rate is defined as:

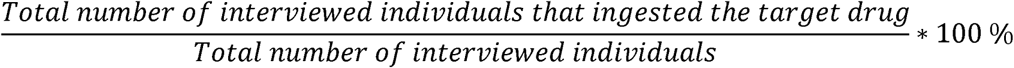

[16].

### Ethical considerations

Ethical permission was obtained from the Federal Ministry of Health. A letter of support outlining the aims and objectives of the survey was submitted from the FMOH to the Regional Health Bureaux and their respective local administrations. Before collecting the data, informed consent was obtained from household heads (HH). In addition, purpose of the survey was explained to HH, and interview was took place only if he or she agreed to sign on the digital consent form on the data collection tablet.

## Results

### Characteristics of study participants

A total of 8,446 SAC were eligible for the study. Out of these, 8154 participated in the study giving a response rate of 96.5%. Out of the interviewed SAC, 1108 were from districts with integrated approach; while 7046 were from districts with vertical approach. In addition, 7,840 (96.15%) and 314 (3.85%) were interviewed during the first and second visits, respectively.

Regarding socio-demographic characteristics of participants: 3939(48.31%) SAC were within age group of 5-9 years; the gender proportions of SAC showed male and female were almost equally involved in the survey; and almost all children (99.7%) were enrolled in primary or secondary (**Table 1**).

**Table 1.**
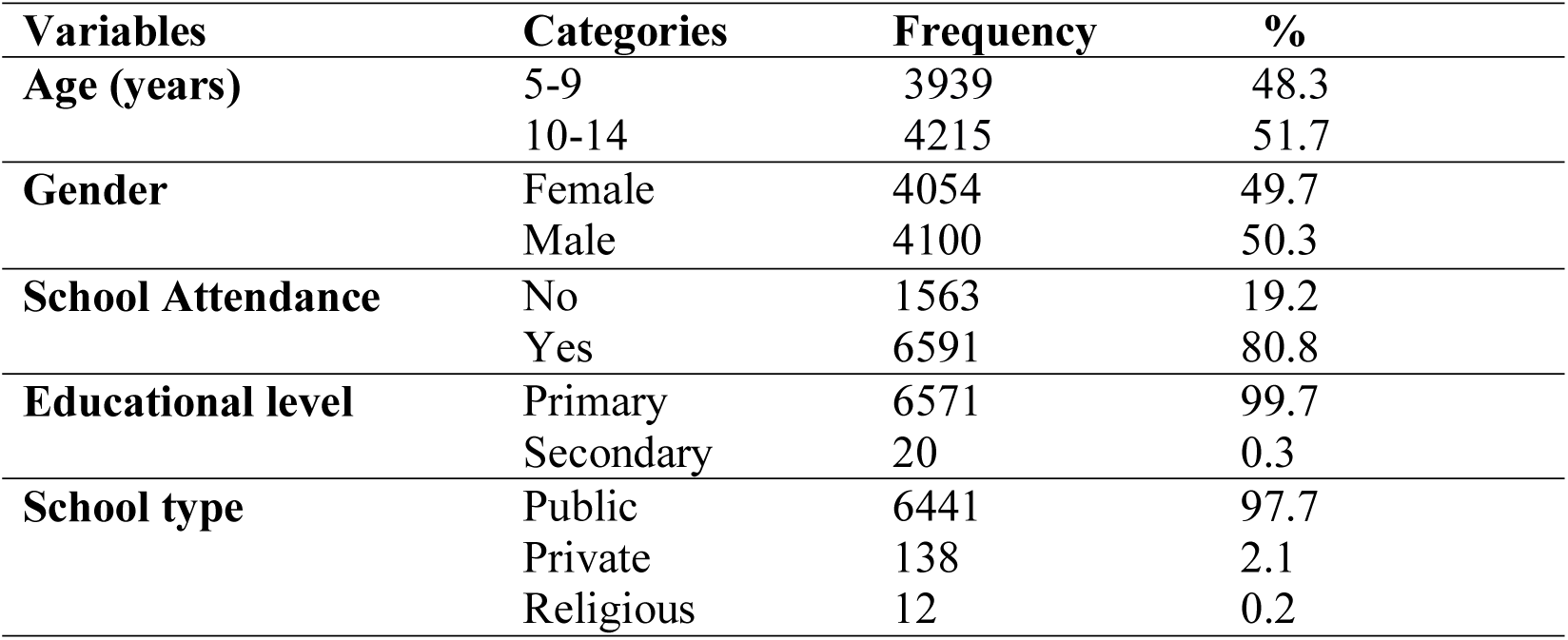
Socio-demographic characteristics of SAC among selected districts of Ethiopia, 2019.

### Albendazole/Mebendazole treatment coverage

The overall treatment coverage of Albendazole/Mebendazole against STH was found to be 71.0% (5785/8154) **(Fig 2)**. Out of 7046 interviewed SAC in districts with non-integrated approach, 4822(68.4%) reported that they swallowed ALB/MBD for STH. Among 1108 interviewed SAC in districts with integrated approach, 963(86.9%) reported that they swallowed ALB/MBD against STH. Dis-aggregated data on treatment coverage of Albendazole by districts showed Guagusa had the highest (91.6%) coverage whereas Gura-Ferda had the lowest coverage (21.6%) (**Fig 2**).

**Fig 2.**
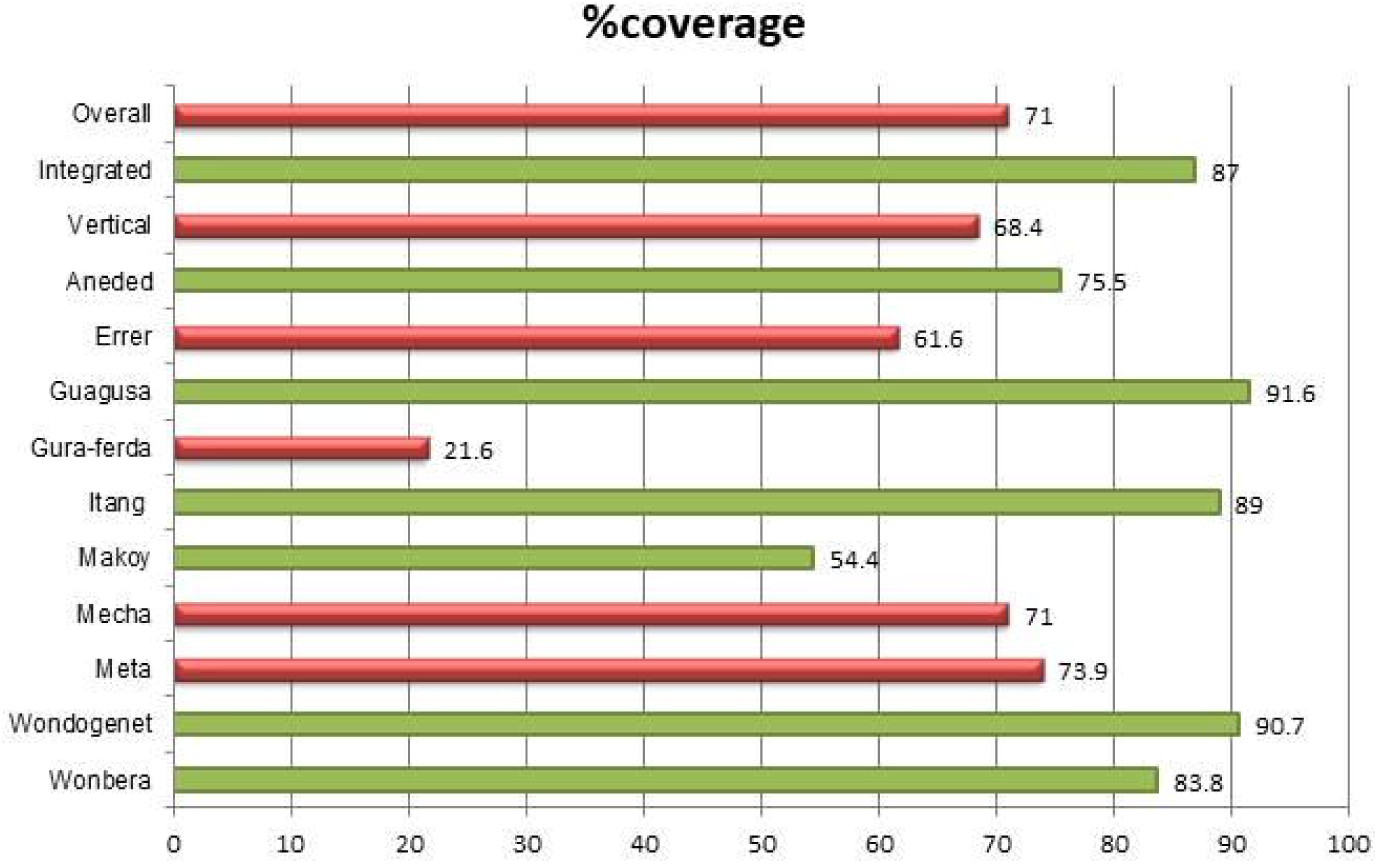
Albendazole/Mebendazole treatment coverage among SAC in Ethiopia, 2019.

### Albendazole/Mebendazole treatment status disaggregated by gender, age and school attendance

The proportion of STH treatment coverage among males was 2948(72%) while among females was 2837(70%). Treatment coverage in the age group (10-14) years was significantly higher than coverage in group (5-9) years old, (x^2^ = 170.57, P-value <0.001). In addition, the treatment coverage in school attendant was higher than coverage non-enrolled children (**Table 2**).

**Table 2.** **Albendazole/Mebendazole treatment coverage disaggregated by age, gender and school attendance in Ethiopia 2019, (N= 8,154).**

| Variables | *Swallowed ALB/MBD* |  |  | Pearson_ chi square<br>(p_ value) |
| --- | --- | --- | --- | --- |
|  | Yes<br>n (%) | No<br>n (%) | Unknown<br>n(%) |  |
| **Gender** |  |  |  |  |
| Female | 2837(70.00) | 1095(27.00) | 122(3.00) | **3.81(0.15)** |
| Male | 2948(71.90) | 1042(25.4) | 110(2.7) |  |
| **Age in years** |  |  |  |  |
| 5-9 | 2534(64.4) | 1246(31.6) | 159(4.0) | **170.57(0.001)** |
| 10-14 | 3251(77.1) | 891(21.2) | 73(1.7) |  |
| **School attendance** |  |  |  |  |
| Yes | 5343(81.1) | 1178(17.9) | 70 (1.0) |  |
| No | 442(28.3) | 959(61.4) | 162(10.3) |  |
| **Heard on MDA** |  |  |  |  |
| Yes | 5227(86.7) | 783(13) | 19(0.3) |  |
| No | 557(26.2) | 1353(63.8) | 213(10.0) |  |

### Reasons for not swallowing ALB against STH

A total of 2137 (26.2%) children reported that they didn’t receive STH treatment. The main reported reasons for not taking the medication were not attending school (n=422, (19.75%) and drugs was not given 397(18.6%) (**Table 3**).

**Table 3.** Reasons for not swallowing Albendazole in selected districts of Ethiopia, 2019.

| **Variables** | **Frequency** | **%** |
| --- | --- | --- |
| *Reasons for not swallowing ALB/MBD* |  |  |
| **Fear of side effects** | 136 | 6.36 |
| **Too sick** | 25 | 1.20 |
| **Not eaten break fast** | 5 | 0.23 |
| **Drug not given** | 397 | 18.60 |
| **No MDA** | 227 | 10.60 |
| **Not attending school** | 422 | 19.75 |
| **Absent at the time of MDA** | 246 | 11.51 |
| **Not living in village** | 52 | 2.42 |
| **Drugs run out** | 75 | 3.51 |
| **No definite reason** | 313 | 14.65 |
| **Bad rumors** | 26 | 1.21 |
| **Un aware about MDA** | 213 | 9.90 |

### Treatment coverage of ALB/MBD in vertical versus integrated treatment approaches

The difference in treatment coverage of ALB/MBD was statistically different between the vertical and integrated approaches (X^2^ = 158.72, P = <0.001) **(Table 5)**.

**Table 5.** Treatment coverage of ALB/MBD in vertical Vs. integrated approaches, Ethiopia, 2019.

| Treatment approach | Swallowed ALB/MBD | | | $X^2$ | P-value |
| --- | --- | --- | --- | --- | --- |
|  | Yes (%) | No (%) | Unknown (%) |  |  |
| Vertical | 4822(68.4) | 2008(28.5) | 216(3.1) | 158.72 | < 0.001 |
| Integrated | 963 (86.9) | 129(11.6) | 16(1.5) |  |  |

### Mass drug administration distribution sites for ALB/MBD

The main site where children received the treatment was in school compound as evidenced by the high percentage value 5223(91%) shown in pie chart below **(Fig 3)**.

**Fig 3.**
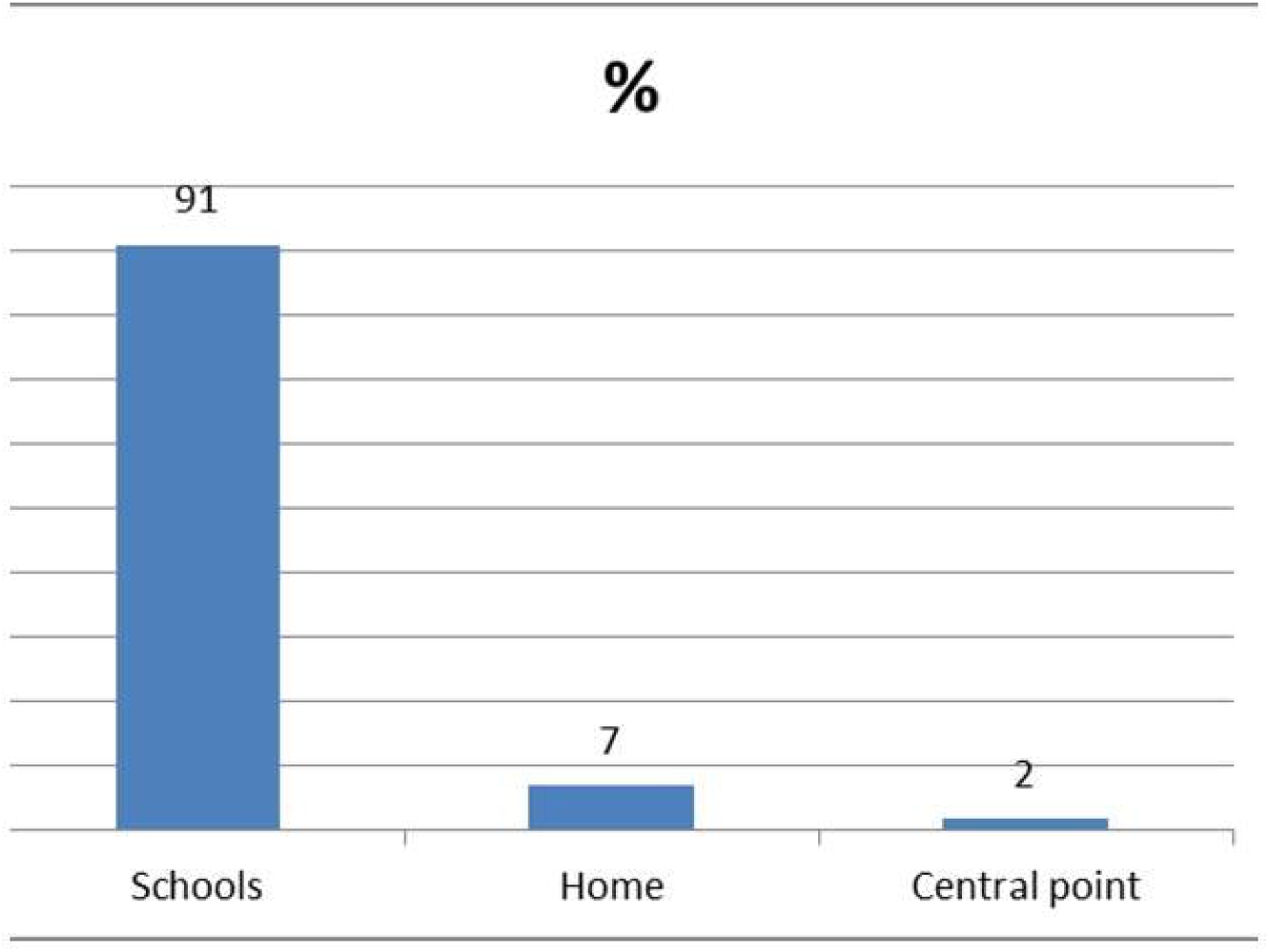
Place of deworming using Albendazole/Mebendazole in Ethiopia, 2019.

## Discussion

This study in ten districts of Ethiopia shows Albendazole/Mebendazole mass treatment against soil-transmitted helminthiases among school-age children in integrated versus vertical treatment approaches result in overall treatment coverage of 71%. In vertical treatment approach 68.4% (4822/7046) were treated; while in integrated approach 86.9 % (963/1108) individuals were treated. The overall treatment coverage in our study is lower than the WHO’s and Ethiopian target; which is to achieve at least 75% coverage of PC by 2020 either through annual or biannual treatment of SAC [9, 19]. The overall coverage in our study is slightly higher than the a global coverage of 68.8% in 2017; which is presented in the WHO fact sheet [15]. In addition, a policy platform on treatment coverage with ALB/MBD of different countries using 2016 WHO data showed 31 countries achieved <75% coverage including Ethiopia, while 57 countries achieved 75% and above treatment coverage [19].

Moreover, WHO report in 2017 showed 36 countries reached 75% coverage for SAC [15]. In our study, the treatment coverage in integrated approach is 87%; it is greater than the WHO recommendation (75%). This result is consistent with a study conducted in Mali where 87% coverage was reported [20]. The possible explanation for higher coverage of ALB/MBD in integrated approach than the vertical approach could be due to its effectiveness to deliver the drugs to those who are in need of drugs at community level.

In our study, gender dis-aggregated data showed the proportion of STH treatment coverage among males was 2948(72%); while among females it was 2837(70%), which means the coverage among males is slightly higher for males. However, this finding dis-agree with a data obtained from 16 countries in which the coverage for females was slightly higher than the coverage for males [21]. Moreover, the coverage of ALB/MBD in this survey is lower than coverage survey of Ethiopia conducted in 2015 [22].

The treatment coverage in school attendant was 81%; which is higher than coverage non-enrolled children (28%). This finding is differ from the coverage survey result of Ethiopia in 2015; which is 92.95% for school attendant while 52.2% for non-enrolled children [22]. The possible explanation for the lower coverage of ALB/MBD among non-enrolled children in this study could be exclusive school based deworming platform is inadequate to address those children who are out of school.

Moreover, our study showed the treatment coverage in the age group 10-14 years was significantly higher than coverage in group 5-9 years. Probably the higher coverage in this age group is due to these children have better chance of attending school. In addition, the main reported reasons for not taking treatment were not attending school and treatment was not given. These reasons can be explained as attending school can increase in association with accessing drugs for those who are in needs. In this case evidence is lacking to discuss over this finding.

The study has the following limitations: First, purposive selection and small of districts make difficult to generalize the findings to national level. Second, identifying barriers associated with low treatment coverage of ALB/MBD among non-enrolled school-age children could be difficult with descriptive cross-sectional study. However, as strength we identified possible reasons for not swallowing the drugs; and this study showed validated treatment coverage of ALB/MBD at national level.

## Conclusions

The study showed Albendazole/Mebendazole mass treatment coverage against soil-transmitted helminths among school-age children was found to be 71%, which is below the WHO recommendation (75%). In addition, coverage survey should be performed in many districts of Ethiopia to validate routine reports. Moreover, operational research is required to identify factors with low coverage of school AlB/MBD among non-enrolled school-age children. Further, a call for action is required from different stakeholders to improve treatment coverage of ALB/MBD.

## Data Availability

All data are available within the manuscript.

## Acknowledgements

Authors would like to thank NTD focal persons in each of the woredas involved in the study; community guiders who were directing households in each segment of the kebeles; study participants, and staff in College of Medicine and Health Sciences who participated in the data collection process.

## Author’s contributions

MA drafted the original manuscript. MA, ZZ, CC, MY, YC and AT contributed in data analysis. MA, ZZ, CC, FS, MY, YC, AT, BG and NN involved in the conception, design & interpretation of the data. All authors read and approved the final manuscript.

